# Machine Learning Analysis of User Sentiments in Tinnitus Management Apps

**DOI:** 10.64898/2026.02.19.26346680

**Authors:** Muhammad Nouman Yousaf, Muhammad Naveed Anwar, Nida Naveed, Umaima Haider

## Abstract

**Background:** Tinnitus affects a substantial proportion of the global population and can severely disrupt sleep, mood, and daily functioning, yet the quality of mobile health apps designed for tinnitus management remains highly variable. Traditional evaluation methods, including clinical trials, expert rating scales, and small-scale surveys, rarely capture large-scale, feature-level feedback from real-world users, leaving a gap in understanding which app characteristics drive sustained engagement and satisfaction.

**Methods:** This study analysed 342,520 English-language reviews from 84 tinnitus-related apps on iOS and Android collected between 2015 and 2025. A pipeline first applied VADER-based preprocessing and sentiment assignment, then trained a graph neural network aspect-based sentiment analysis (GNN-ABSA) model operating on sentence-level dependency graphs to infer feature-level sentiment for domains such as sound therapy, sleep support, pricing, advertisements, stability, and user interface.

**Results:** The GNN-ABSA model achieved an accuracy of 84.4% and a macro F1 score of 0.829 on unseen aspect-level test data, indicating stable performance across sentiment classes. Therapeutic features like sound masking and sleep support were associated with predominantly positive sentiment, whereas pricing, advertisements, background playback, and technical stability attracted more neutral or negative feedback over the ten-year period.

**Conclusions:** Large-scale, graph-based feature-level sentiment analysis provides a user-cantered perspective that complements clinical trials and expert app quality ratings, offering actionable guidance for developers seeking to prioritize design improvements, supporting clinicians in recommending suitable apps to patients, and informing the design of more explainable and user-driven digital health tools.

**Trial Registration:** Not applicable. This study analysed publicly available app store reviews and did not involve human participants.

## 1. Background

Tinnitus is a common auditory condition in which people perceive sounds such as buzzing or ringing without any external sound source. Tinnitus affects approximately 10-15% of the global population, and for many, it is a serious, lifelong condition that causes psychological issues including anxiety, depression, and poor sleep quality. Due to limited clinical availability, traditional evidence-based treatments like Tinnitus Retraining Therapy (TRT) and Cognitive Behavioural Therapy (CBT) are expensive, time-consuming, and challenging to access.

Mobile Health (mHealth) apps have emerged as a practical solution for providing management tools, such as sound therapy and online counselling, directly to users. However, the application market is currently fragmented and inconsistent in quality. Existing research tends to focus either on clinical outcomes reported in controlled trials or on expert-driven quality assessments such as the Mobile App Rating Scale (MARS). These approaches do not fully capture user dissatisfaction or the specific design issues that lead to reduced engagement or app abandonment in everyday use.

### 1.1 Current Approaches to App Quality Assessment and the Research Gap

The leading tool for objective mobile app quality assessment is the Mobile App Rating Scale (MARS) [12], [16]. This 23-item instrument measures an app’s functionality, engagement, aesthetics, and information quality. In the first systematic review of its kind, [12] used MARS to evaluate 60 tinnitus apps (34 Android, 26 iOS), which yielded average scores from 2.65 to 4.60 out of 5. A crucial finding was that all of the widely available apps lacked any scientific evidence for their effectiveness.

Building on this, [16] took a two-part approach. First, they surveyed 643 people with tinnitus and discovered that 75% had never used a tinnitus app. Then, they used the MARS tool to evaluate 18 available apps, finding their quality scores ranged from 1.6 to 4.2, with sound therapy being the most common feature. Expert evaluations like MARS are limited by their objectivity. They fail to track the subjective frustrations of real users or the subtle design flaws that ultimately lead to app abandonment.

### 1.2 Bibliometric and Systematic Review Findings

In a [7] study, performed a broad analysis of mobile health (mHealth) research for tinnitus, reviewing 75 publications and 28 apps from 15 different countries. Their work showed that Germany produced the most research, accounting for 25% of the publications. More importantly, they found that about 79% of the available apps had no scientific studies to prove they worked. This clearly highlights a major gap: while there are many apps on the market, very few are developed with solid evidence.

While these systematic approaches have comprehensively catalogued available apps and their technical characteristics, they do not address user-perceived effectiveness, satisfaction, or the specific features driving user satisfaction or frustration in real-world conditions.

### 1.3 Identification of Research Gap

Key Limitation of Prior Research: Previous studies on tinnitus apps employed either:

- Clinical outcome measures (THI, TFI, anxiety/depression scales) on small trial samples (42-84 participants) [20], [17], [18].
- Expert quality ratings (MARS) requiring manual evaluation [12], [16].
- Web-based surveys on limited populations (643 respondents) [16].
- Qualitative thematic analysis of focus groups [11].

None of these approaches:

- Analysed sentiment from large-scale user-generated app reviews.
- Performed feature-level sentiment analysis (aspect-based sentiment analysis).
- Applied advanced natural language processing or machine learning
- Captured user perspectives at scale (>100,000 reviews)
- Provided temporal trend analysis (10-year coverage)

This gap represents a significant opportunity for computational analysis of user-generated content to complement traditional clinical and expert-driven methodologies.

### 1.4 Data Selection Rationale

The dataset selection criteria were deliberately designed to maximise analytical power, diversity, and real-world relevance. The ten-year period from 2015 to 2025 was selected to provide comprehensive coverage of mHealth app development and evolution. This timeframe encompasses:

- **Early mHealth adoption**: Initial smartphone-based tinnitus interventions and early user feedback
- **Growth and refinement**: Increased app availability and evolving feature sets
- Maturation and AI integration (2020-2025): Introduction of machine learning features, advanced sound algorithms, and integration with wearable devices

Studying the last decade offers a valuable perspective. This timeframe is ideal because it is recent enough to be applicable to today’s mobile health standards, yet long enough to identify clear, long-term patterns in user sentiment and feature development.

### 1.5 App Coverage and Inclusion Criteria

Rather than attempting exhaustive coverage of all available apps, the study employs strategic sampling of 84 applications selected to ensure diversity and relevance. Inclusion criteria encompass:

- Dedicated tinnitus management apps (e.g., ReSound Relief, calm, mynoise [28])
- Hearing and audiology applications with tinnitus management features
- General wellness apps (meditation, sleep, sound masking) used by tinnitus patients

This inclusive approach reflects ecological reality: many tinnitus sufferers use multiple app types—dedicated tinnitus apps alongside general wellness, sleep, and meditation applications—for comprehensive symptom management [16]. This broader coverage enhances the study’s real-world validity and captures the diverse strategies users employ for tinnitus management.

### 1.6 The Case for Scalable Sentiment Mining

User reviews on app stores include a wide range of detailed feedback that can be difficult to analyse using traditional qualitative methods. Common features of these reviews include:

- Short, informal text: Reviews average 50-150 words with informal language and abbreviations
- Multiple, sometimes conflicting sentiments: Individual reviews frequently express contradictory assessments (e.g., “great sound quality but terrible user interface”)
- Domain-specific terminology: References to technical features, hearing aid compatibility, app stability
- Non-standard syntax: Informal punctuation, emoji usage, phonetic spelling
- Noise and spam: Some reviews unrelated to the app’s core function

Manually analysing detailed feedback linked to specific app features isn’t practical for a dataset of over 342,000 reviews. Automated methods are needed to find patterns between app features and user opinions on such a large scale.

### 1.7 Technique Used in Prior Tinnitus App Research

Table 1 summarizes the methodologies employed in prior tinnitus app research, highlighting their sample sizes, strengths, and key limitations.

**Table 1.**
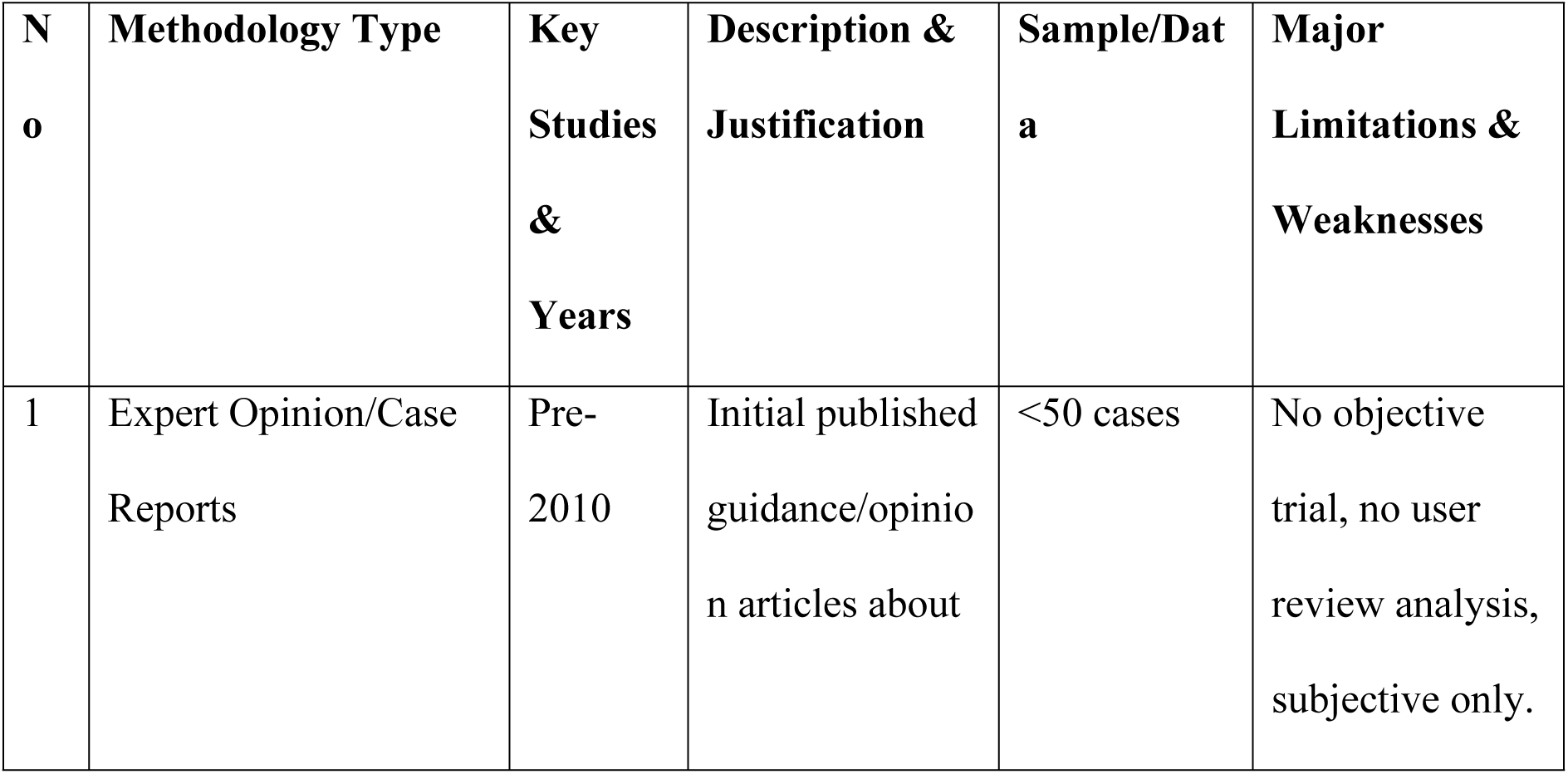
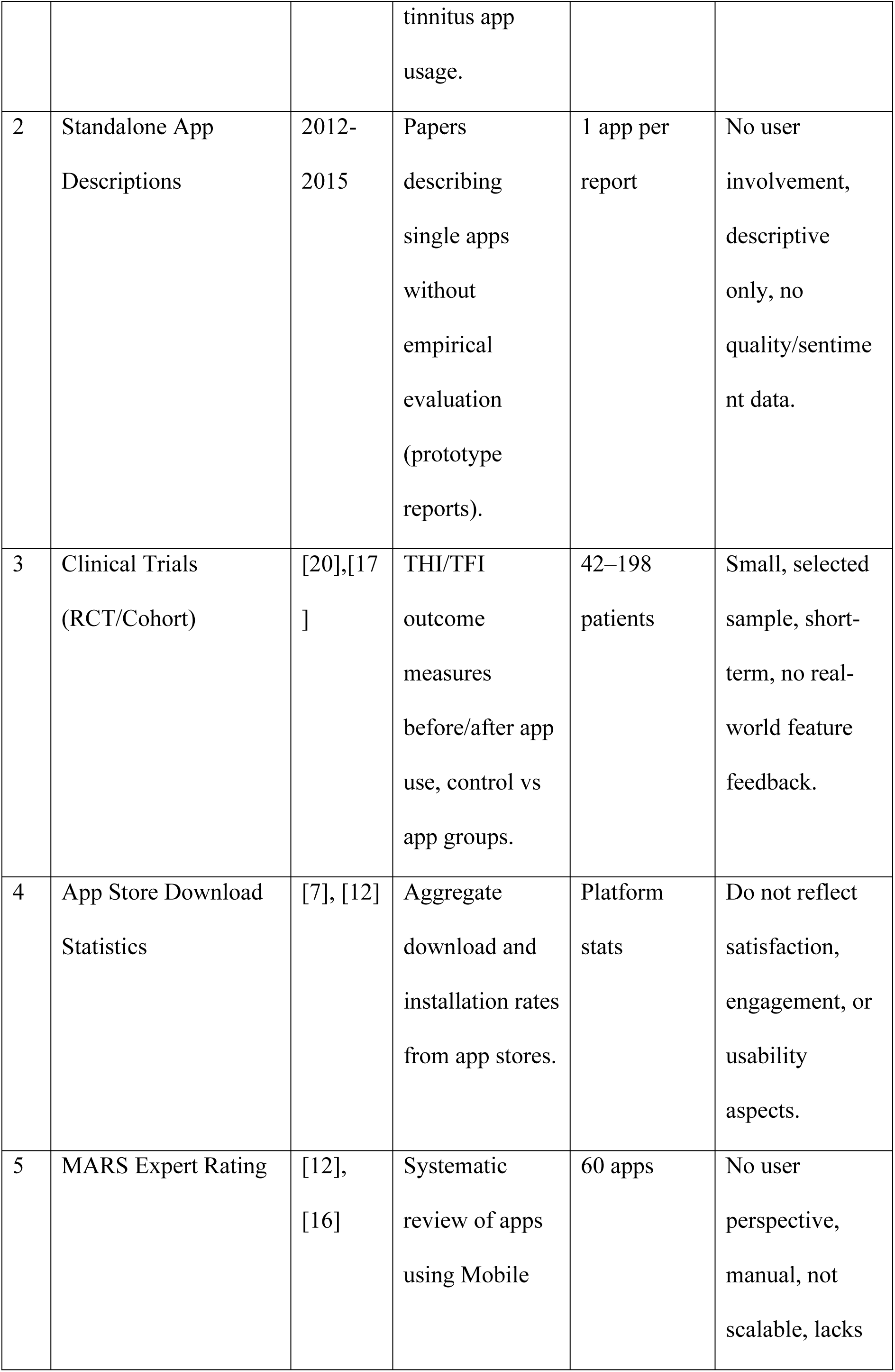
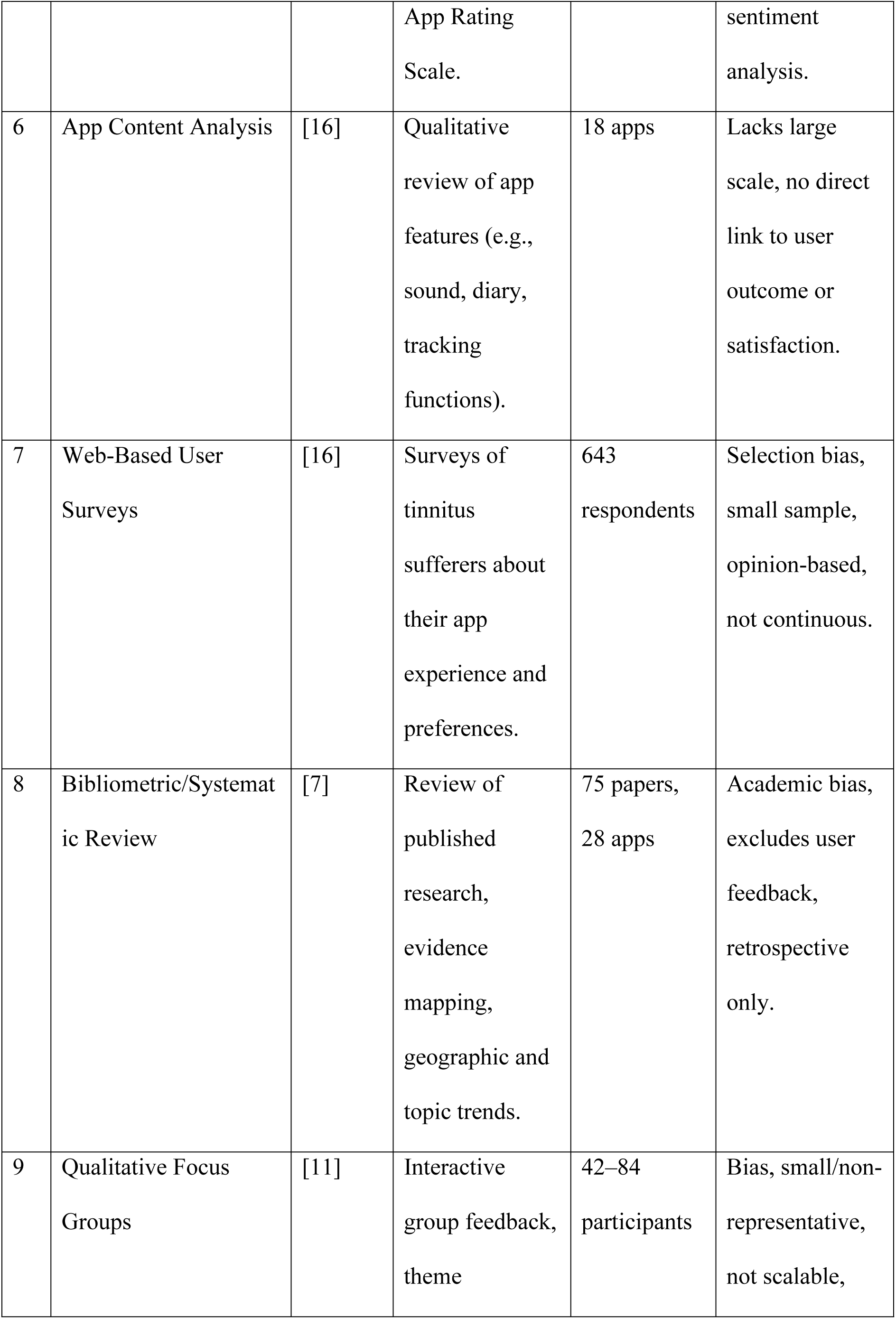
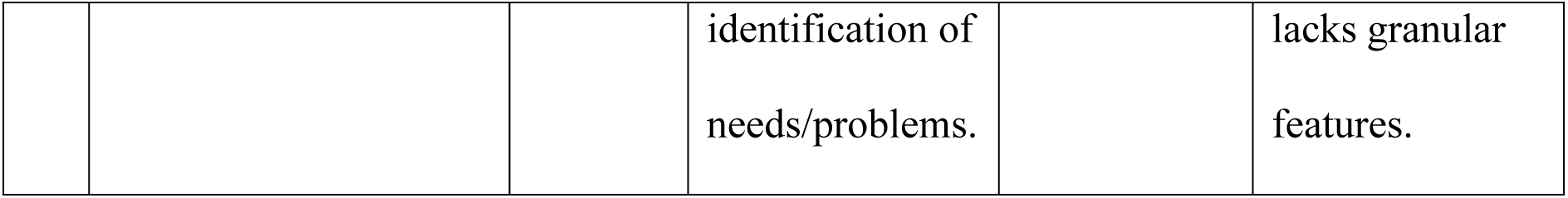
Prior Tinnitus App Research Methodologies and Limitations.

1. Expert Opinion and Case Reports (Pre-2010): Initial research into tinnitus apps was largely based on expert opinions and individual case studies. While this helped form a basic understanding of how apps could be used, the methods lacked scientific rigor, objective data, and input from actual users. Although expert insight is useful for forming initial ideas, it cannot be applied to the broader population and does not represent the real-world experiences of users on a large scale [7].

2. Standalone App Descriptions (2012–2015): With the rise of mobile tech, researchers began documenting their own tinnitus apps. However, these early papers were often just descriptive; they focused on features and intended functions while ignoring essential user testing, clinical trials, or comparisons with existing treatments. While this kind of descriptive work is a useful starting point, it tells us nothing about whether the apps actually helped users or improved health outcomes [12].

3. Clinical Trials Using Outcome Measures (2015–2024): Recent randomized trials and cohort studies (like [20], [17]) mark a major step forward in tinnitus app research. By using standardized measures like the THI and TFI, they have proven that some apps can significantly reduce symptoms. However, these studies have inherent limitations. They are conducted under tightly controlled conditions with small, specific groups of participants (usually between 42–198 people) over short periods like 3 to 6 months. Their focus is solely on whether symptoms improve on average. Because of this narrow focus, a critical question remains unanswered: which specific app features make users want to keep using them? These trials do not tell us what drives user satisfaction, engagement, or real-world adoption—the very factors that determine if an app will be successful outside of a research setting [18].

4. App Store Download and Installation Statistics (2015–2020): Download counts show what’s popular, but they don’t tell the whole story. Because marketing and app store rankings often drive downloads more than actual quality, these numbers fail to show if people are actually using the app or which features are frustrating them [7].

5. Mobile App Rating Scale (MARS) Expert Evaluation (2019–2020): The MARS system was basically the first standardized method for checking app quality, where they get a panel of experts to judge apps on things like how they function, their design, if they’re engaging, and the information quality. Studies have shown MARS has good reliability between different raters (κ = 0.82–0.94) and it’s been used to compare and rank different apps that are out there ([12], [16]. But the problem is, MARS is all from an expert’s point of view, not a regular user’s. The things a expert looks for might be totally different than what a user cares about, and they can easily miss usability problems that really frustrate people actually using the app. Also, this method can’t really handle the hundreds of apps in the app stores today [12]. And most important, the scores from MARS don’t actually line up with how satisfied users say they are in their reviews and ratings [16].

6. App Content Feature Analysis (2019–2020): When you look at what features tinnitus apps have—like if they offer sound therapy or symptom tracking, hearing aid integration, or community support—it tells you what’s out there. This kind of research showed that sound therapy is in almost all of the apps, and that most of them don’t really have content that’s based on solid evidence. But just because a feature is there, doesn’t mean it’s any good. It doesn’t tell you if users are happy with how it works, or what features they actually find useful in their day-to-day life [16].

7. Web-Based User Surveys (2019): So when they do surveys of people with tinnitus—like that one with 643 people [16]—you get some really good info straight from the users about what they know and what’s stopping them from using apps. That’s how they found out that a huge 75% of people had never even tried a tinnitus app, which really shows you there’s a big adoption problem. But surveys have their issues too. The people who answer them are kind a self-selected so they might be different than people who don’t, the sample size is still pretty small when you think about how many people have tinnitus worldwide, and it’s just a snapshot in time. The opinions you get from surveys just aren’t detailed enough to tell you exactly which features people love or which ones makes them frustrated [16].

8. Bibliometric and Systematic Literature Review (2020–2023): Large-scale reviews are excellent for spotting industry trends. For instance, [7] analysed 75 papers and discovered that a staggering 79% of tinnitus apps lack scientific backing. This highlights a major gap between app availability and actual evidence. However, these reviews suffer from publication bias, as unsuccessful trials rarely make it into journals. Furthermore, they overlook a massive data source: the raw feedback and frustrations found in app store user reviews [7].

9. Qualitative Focus Group Discussions (2022): Focus groups are difficult and costly to organise, and typically involve only a small number of participants who are unlikely to reflect the wider user base. In many cases, a few dominant voices can shape the whole discussion, which limits how representative the findings are. As a result, the output is mainly narrative—stories and quotations—without the kind of large-scale quantitative evidence that can show what really matters to the many users leaving app store reviews [11].

### 1.8 Graph Neural Network for Aspect-Based Sentiment Analysis

Researchers are increasingly moving toward Graph Neural Networks (GNNs) to make sense of the messiness in user reviews. Older models tended to view text as just a flat string of words, but GNNs actually map out how words connect and relate to one another ([21], [22]). This shift is a big deal for sentiment analysis; it allows us to pinpoint exactly which product features a user is mentioning and, more importantly, how they feel about those specific parts rather than just the whole app.

### 1.9 Core Advantages of GNN-ABSA

A key strength of graph models is their ability to represent text as a connected network. Each word becomes a node, and grammatical relationships between words form edges. This structure captures long-range dependencies in language—context that traditional sequential models often struggle to identify [19]. Think about a phrase like ‘NOT effective.’ A simple model might just see the word ‘effective’ and get the sentiment wrong, but a dependency graph links the negation directly to the adjective, stopping that error before it happens.

The main advantage of GNN-ABSA is its ability to analyse complex user feedback. Tinnitus patients often mention several issues in a single comment, for example: “The sound quality is excellent, but the app crashes frequently and the subscription is too expensive.” A simple model may struggle with such mixed opinions, but this method separates each sentiment clearly—recognising the positive view of sound quality while identifying problems with reliability and cost. This level of analysis goes beyond basic rating systems, providing developers with specific insights into areas that require improvement [22], [21].

A notable advantage of graph models is their interpretability. Unlike many traditional models that operate with limited transparency, graph models make it possible to trace how conclusions are formed. For example, by examining the attention weights in a Graph Attention Network (GAT), it becomes clear which words or relationships had the strongest influence on a given prediction [8]. This level of clarity is particularly important in healthcare settings, where clinicians and developers need to understand the reasoning behind a model’s output before they can rely on it.

Transformer models such as BERT and RoBERTa achieve high levels of accuracy but often require substantial computational resources. Recent hybrid methods, including the approach outlined by [1], combine a more compact model like DistilRoBERTa with Graph Neural Networks (GNNs) and still attain around 80–85% accuracy across more than 500,000 reviews. This balance between performance and resource use is particularly beneficial when analysing large datasets, such as the 342,520 tinnitus app reviews used in this study.

### 1.10 Graph Neural Network as Semantic Bridge

Using Graph Neural Networks [23] offers a relatively new but well-tested way to link what users say about app features with the emotions they express. In this study, GNN-based sentiment analysis is useful for several reasons. First, it can connect emotion words directly to particular features, so sentiment can be assigned at feature level. Second, by modelling grammatical structure, it makes the model’s reasoning easier to interpret, because it is clearer why a certain sentiment was linked to a given feature. Third, it can handle large datasets, such as the more than 342,520 reviews used here, without a major loss in performance. Finally, recent studies in digital health [10], [5] show that GNNs work well in healthcare settings, which supports their use in this context.

### 1.11 Novelty and Research Contribution

Although aspect-based sentiment analysis with GNNs has already been used in areas like restaurant and product reviews, this is the first time it has been applied at scale to tinnitus management apps, using more than 342,520 reviews collected over ten years. This helps to fill an important gap. First, it works at a much larger scale than earlier studies, which relied on small surveys or clinical trials. Second, it uses NLP and machine learning instead of manual questionnaires, so the analysis is automated and consistent. Third, it looks at changes over a full decade rather than at a single moment in time. Fourth, it captures what real users say in public reviews, rather than only relying on clinical outcome scores. Finally, it identifies sentiment for specific app features instead of giving just one overall satisfaction score. Together, these points mean the study offers evidence-based, user-focused insights that sit alongside, and add to, traditional expert and clinical evaluations.

### 1.12 Research Questions

This study addresses three core research questions:

RQ1: How is user sentiment distributed across reviews for tinnitus management apps over the period 2015-2025?

RQ2: Which specific app features—such as sound quality, stability, pricing, and usability—dominate user discussions and sentiment patterns in the review data?

RQ3: Can a Graph Neural Network-based aspect-level sentiment analysis (GNN-ABSA) approach improve the precision of sentiment attribution to specific app features compared to document-level approaches?

## 2. Methods

### 2.1 Data Collection

App store reviews were collected using custom Python scripts executed in Google Colab notebooks. Two separate scrapers were developed:

- Apple App Store scraper: Utilized the iTunes RSS feed API to retrieve reviews from iOS applications
- Google Play scraper: Implemented using the google-play-scraper Python library (version 1.2.7) to access Android application reviews

The dataset targeted 84 tinnitus-related mobile applications identified through systematic searches of both platforms, including apps recommended by NHS UK and other prominent hearing health organizations. Data collection was conducted between November and December 2025.

To ensure comprehensive coverage of English-language reviews, both scrapers queried reviews across 15 English-speaking regions:

United States, United Kingdom, Canada, Australia, Ireland, New Zealand, India, Singapore, South Africa, Philippines, Nigeria, Kenya, Malaysia, Hong Kong, and United Arab Emirates.

For each application, the scraper queried all 15 regional app stores separately. This approach was necessary because:

- Regional app stores maintain separate review databases.
- Some reviews are only visible in specific regional stores.
- Complete international coverage required querying each region independently.

The scrapers were configured to retrieve reviews posted between 2015 to 2025, capturing the full historical record available on both platforms at the time of collection.

The scraping process was executed sequentially, with each of the 84 applications processed individually. For each app:

- The appropriate app ID or package name was configured in the scraper.
- The script queried the app across all 15 regional stores.
- Results were saved to a separate CSV file.
- The process was repeated for the next application.

This resulted in 84 individual CSV files containing the raw scraped data.

All 84 individual CSV files were combined into a single master dataset using Python pandas library in Google Colab. The consolidation code merged the files while preserving all original fields (author, date, comment, rating, app name, and platform).

### 2.2 Data Deduplication and Text Preprocessing

A multi-stage deduplication process was implemented to identify and remove redundant entries:

Reviews with identical review IDs, timestamps, and text content were identified as exact duplicates. This stage removed entries where the same review was collected multiple times from different regional queries. Reviews with greater than 95% text similarity (calculated using Jaccard similarity on tokenized text) from the same user on the same app were flagged as near-duplicates. This captured cases where minor formatting differences existed between regional versions of the same review.

For apps available on both iOS and Android, reviews posted by users across both platforms were examined for duplicates, though this was rare.

The deduplication and quality filtering process resulted in a final dataset of 342,520 unique, valid reviews for analysis.

Table 2 summarizes the deduplication process stages and resulting dataset sizes.

**Table 2.** Summary of Data Collection and Deduplication Process.

| Processing Stage | Entry Counts | Percentage Remaining |
| --- | --- | --- |
| Initial Collection (84 apps, 15 regions each) | 5,488,498 | 100% |
| After Exact Deduplication | ~2,100,000 | 38.3% |
| After Near-Duplicate Removal | ~800,000 | 14.6% |
| After Cross-Platform Consolidation | ~400,000 | 7.3% |
| After Quality Filtering | 342,520 | 6.2% |
| Total Duplicates Removed | 5,145,978 | 93.8% |

The high duplication rate (93.8%) is characteristic of multi-region scraping methodologies where the same content is retrieved across multiple geographic API endpoints. This approach, while generating substantial initial volume, ensures comprehensive coverage of all reviews regardless of their regional visibility, at the cost of requiring extensive deduplication processing.

A second preprocessing stage prepared the reviews for modeling. All comment text was converted to lowercase, URLs and emojis were removed using regular expressions, and extra whitespace was stripped from the beginning and end of each review. An intermediate dataset was created containing only the Comment and Rating columns to streamline subsequent processing steps. Apostrophes were normalized and common contractions were expanded using the contractions library, before the text was tokenized and lemmatized. A custom stopword list that preserved important negation words was applied, and the resulting token lists were stored together with the cleaned comment text and corresponding rating values. This preprocessed dataset served as input for the subsequent modeling steps.

### 2.3 Exploratory Data Analysis

An exploratory data analysis (EDA) was conducted on the full tinnitus review dataset to understand its overall structure, quality, and temporal distribution. The combined reviews were loaded into pandas, the Date column was converted to datetime format, and basic statistics were calculated including total review counts, reviews per app, per platform, per year, and the distribution of 1-5 star ratings. Visualizations were created, including bar charts of the ten most-reviewed apps, a yearly line plot of review counts, and a histogram of review lengths. Data quality checks revealed few missing values in the Comment and Author fields but identified a large number of duplicate reviews. Following the removal of 5,145,978 duplicate rows, 342,520 unique reviews remained as the cleaned dataset for subsequent analysis. Figure 1 shows the top 10 apps with the most reviews.

**Figure 1.**
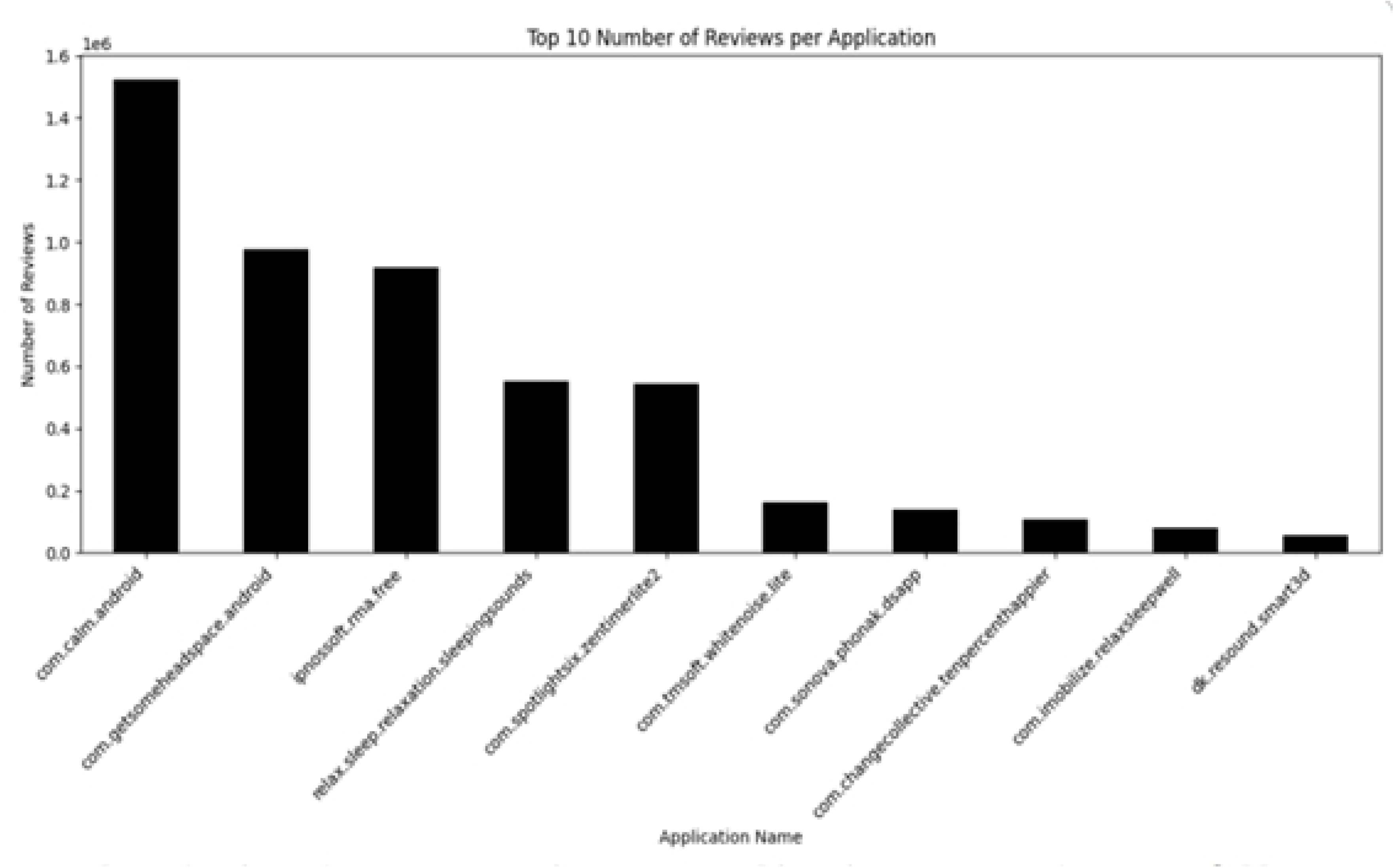
Distribution of user reviews across the top 10 most-reviewed tinnitus management applications. The bar chart displays review counts for apps with the highest number of user reviews in the dataset (n = 342,520 total reviews).

### 2.4 Sentence Splitting

A sentence-splitting step was performed to prepare the cleaned reviews for graph-based, aspect-level sentiment analysis. The en_core_web_sm spaCy model was used to segment each review into individual sentences while preserving the original star rating. For each review, a spaCy document was constructed, sentences of at least three characters were extracted, and a punctuation-based fallback handled very short or unusual text fragments to minimize data loss. Each sentence was stored with metadata including its source file, original row index, position within the review, and rating. This processing stage transformed 342,520 reviews into 785,184 individual sentences, which served as the basic units for constructing the sentence-level dependency graphs used by the GNN model. Figure 2 shows the sentence extraction results.

**Figure 2.**
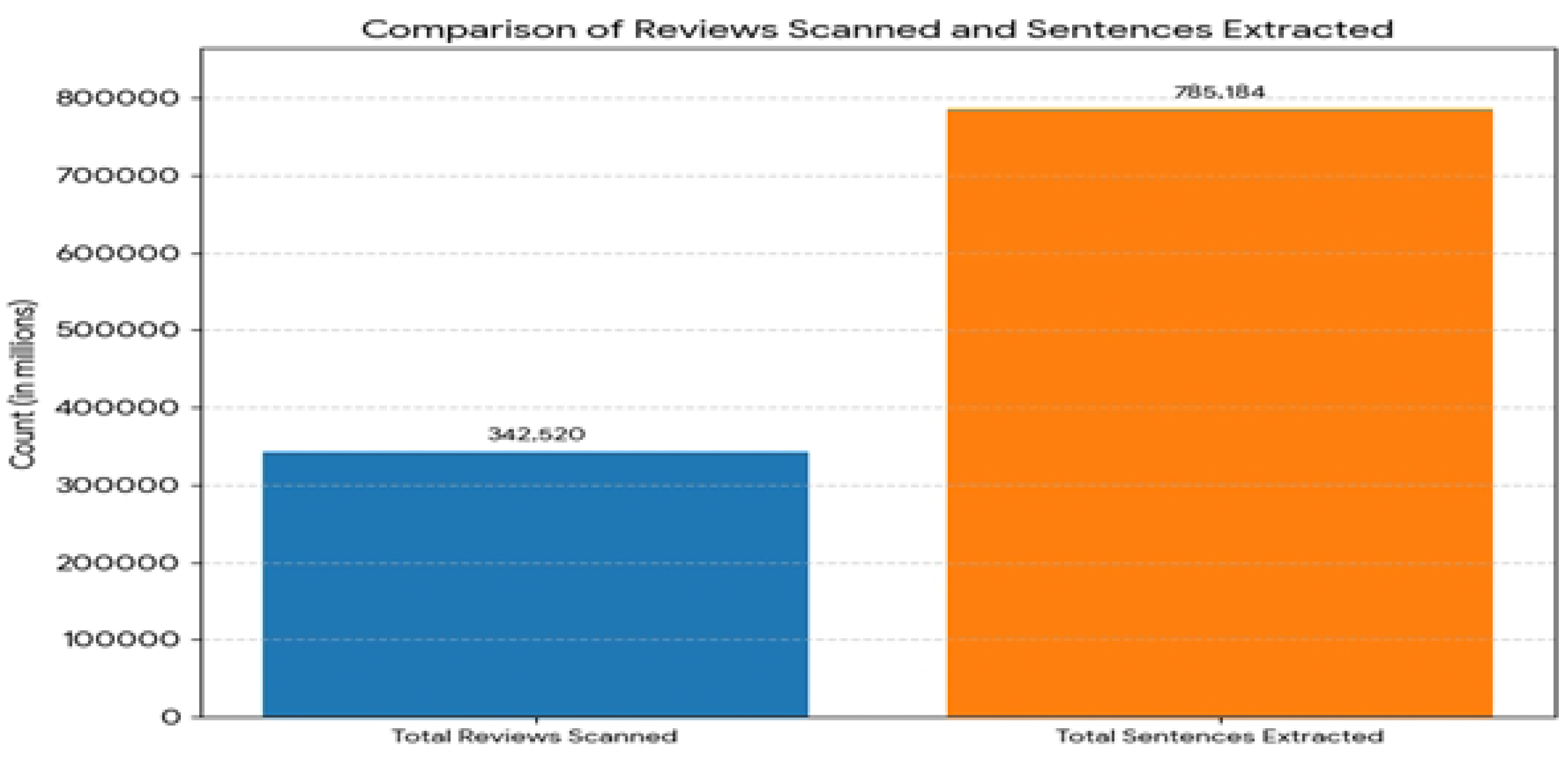
Sentence extraction process from user reviews. The visualization shows the distribution of sentences extracted from the 342,520 reviews, resulting in 785,184 individual sentences for aspect-level analysis.

### 2.5 Aspect detection

A rule-based aspect detection step was applied to identify which app features each sentence referred to. The sentence-level dataset was processed using a predefined set of aspect categories including sound quality, sound variety and personalization, sleep support, usability and interface, stability and performance, battery use and background playback, price and advertisements, developer support and updates, lack of effectiveness, and emotional relief.

Each category was associated with a list of keywords and phrases derived from domain knowledge and preliminary topic analysis. Using spaCy’s PhraseMatcher, each sentence was scanned for these terms, and matched aspects along with their trigger phrases were stored in additional columns, resulting in sentences tagged with their star rating and one or more detected aspects. Binary indicator columns and an aspect frequency summary were generated to quantify how often each aspect appeared across the 785,184 sentences, facilitating subsequent graph construction and feature-level analysis. Figure 3 shows the aspect distribution.

**Figure 3.**
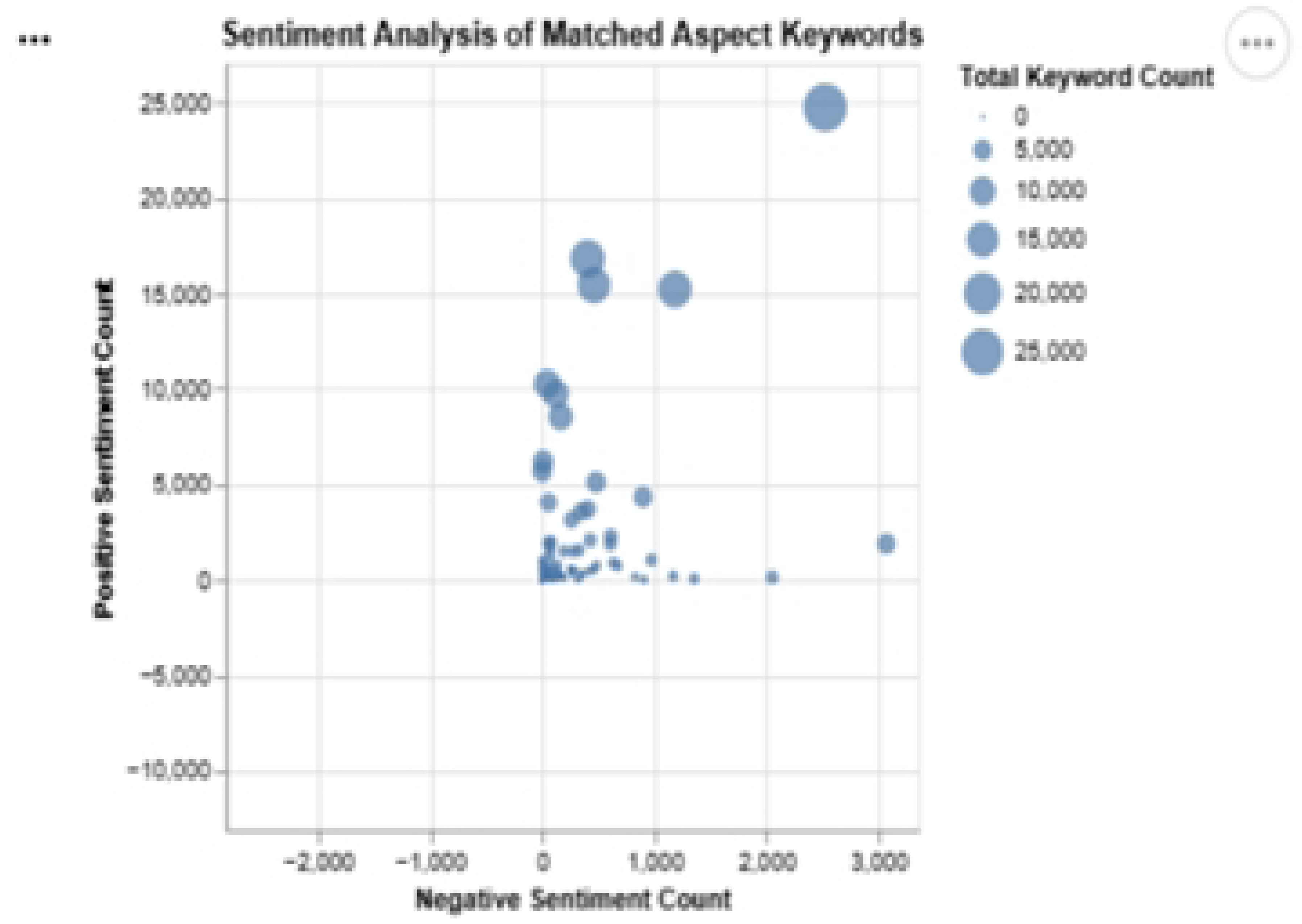
Distribution of aspect categories across detected mentions. The bar chart shows the frequency of different app feature aspects mentioned in user reviews, with pricing and advertisements being among the most frequently discussed categories.

#### 2.5.1 Aspect Keyword Identification and Sources

The aspect keywords for the sentiment analysis were chosen using a mixed approach that drew on earlier research, app feature descriptions, and an initial review of user comments. First, tinnitus-related clinical features such as sound therapy, masking, relaxation, sleep support, and cognitive behavioural therapy were taken from existing studies and reviews of tinnitus and mobile health apps [12], [16], [7]. These works repeatedly describe the main functions found in tinnitus apps and were used as a starting point for defining aspects.

Technical and usability features such as ease of use, performance, adverts, subscription cost, and personalisation were added based on previous app quality studies and large-scale sentiment work on mobile health tools [16], [5]. To check that these aspects matched what real users talk about, common words and phrases were explored in the collected app store reviews during the exploratory analysis stage. This helped adjust the aspect list so that it followed user language while still staying close to the research background. The final aspect set was therefore built gradually, using both the literature and patterns in the data, rather than copied from a single source.

### 2.6 Sentiment Assignment

A third preprocessing step added sentiment labels to each detected aspect in the sentences using a rule-guided VADER pipeline in Google Colab. VADER (Valence Aware Dictionary and sEntiment Reasoner) is a lexicon and rule-based sentiment analysis tool specifically designed for social media text and user-generated content [9]. It was selected for this study because it handles informal language, punctuation-based emphasis, and negation patterns commonly found in app store reviews. The sentiment assignment process involved the following steps:

1. **Context Window Extraction**: For every sentence containing one or more detected aspects, spaCy was used to identify words in the local context around each aspect term.
2. **Opinion Word Detection**: Nearby opinion-bearing words were extracted, including adjectives (e.g., “excellent”, “terrible”), adverbs (e.g., “very”, “barely”), and sentiment-bearing verbs (e.g., “love”, “hate”). Additional rules captured domain-specific negative terms such as “crash”, “bug”, “freezes”, and “stops working”.
3. **VADER Sentiment Scoring**: The extracted opinion words were joined into a short text span and passed to the VADER sentiment analyser [9]. VADER computed a compound sentiment score ranging from −1 (most negative) to +1 (most positive) for each aspect mention.

This process produced 357,866 aspect-level sentiment labels from 785,184 sentences. Summary statistics were generated showing, for each aspect category, the distribution of positive, negative, and neutral sentiment mentions. These VADER-derived labels served as the training targets for the subsequent GNN-based sentiment classification model.

Figure 4 shows the aspect keywords

**Figure 4.**
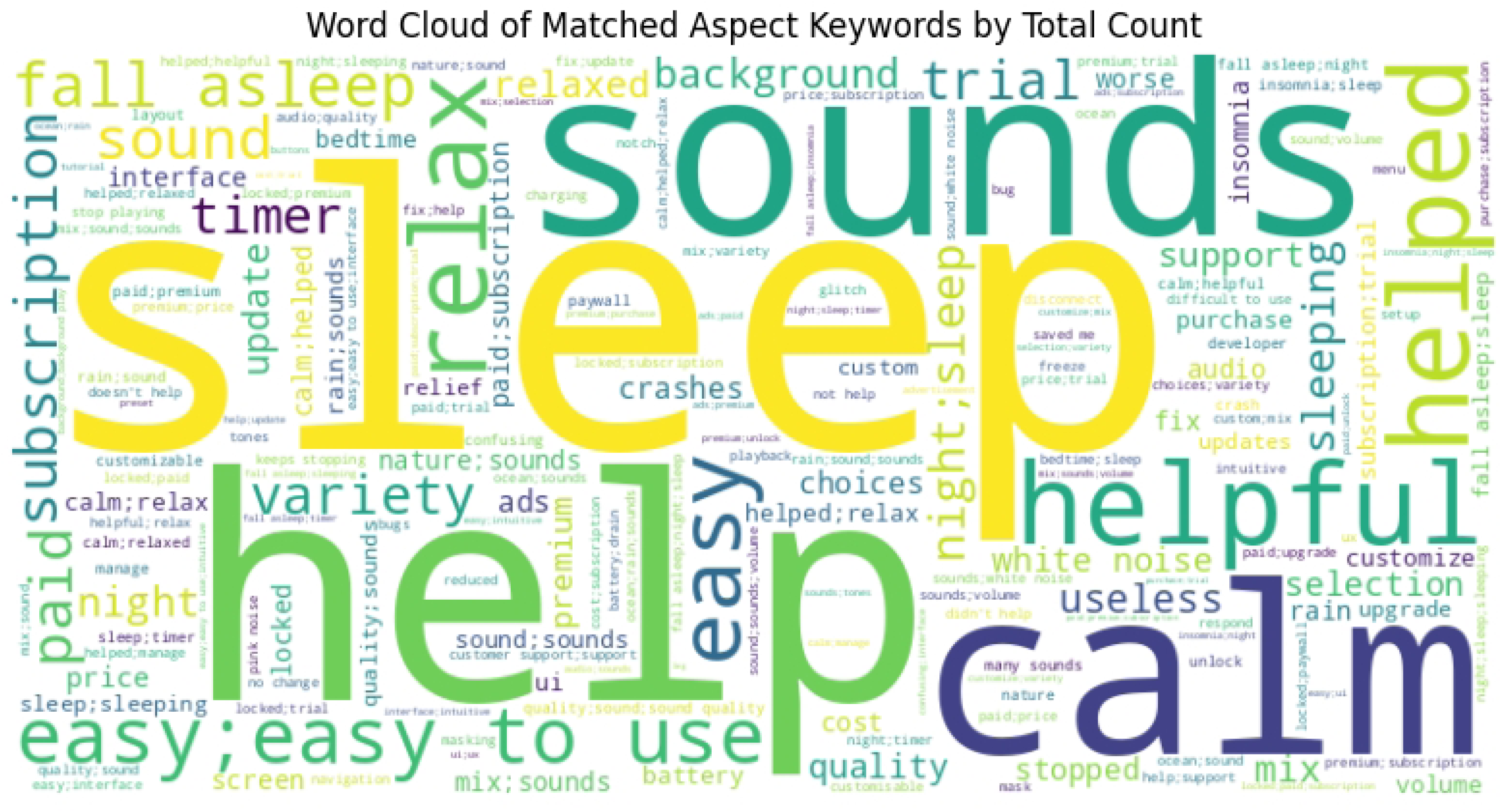
Word cloud visualization of aspect-specific keywords. The visualization displays the most common terms associated with each aspect category identified through the aspect detection process.

### 2.7 Experimental Setup

Here are the some facts about Experimental setup

- The sentence-level preprocessing step produced 357,866 separate graphs, with each graph representing one sentence–aspect pair.
- Of these, 300,000 graphs were used for training, and the remaining 57,866 graphs were kept aside as unseen data for validation and testing.
- All experiments were run on a local machine using only a CPU, without any GPU or other specialised hardware.
- Model performance was tracked using overall accuracy, macro-averaged F1 score, and confusion matrices for the three sentiment classes (positive, negative, and neutral).

### 2.8 GNN Configuration

The sentiment model was implemented as a Graph Attention Network (GAT) using the PyTorch Geometric library. Each input example was a sentence converted into a small graph, which then passed through two GAT layers in sequence. In the first layer, multiple attention heads built a hidden representation for each node, and the second layer refined this so that nodes could take more of their local context into account. Both layers used four attention heads, and the total hidden size was set to 128.

The node features came from the earlier preprocessing steps and were stored in each graph as feature vectors for every node. The size of these vectors was taken from the data and kept fixed across all runs. After the graph layers, an aspect mask was applied so that only the nodes linked to the detected aspect terms were used for the final prediction. This meant the model focused on the part of the sentence that described the aspect, rather than on the whole sentence.

Once the aspect nodes were selected, a dropout layer with a rate of 0.3 was applied to reduce overfitting, and a fully connected layer then mapped the hidden features to three output classes: negative, neutral, and positive sentiment. The model was trained with cross-entropy loss. Training used the Adam optimiser with a learning rate of 1×10-4, mini-batches of 16 graphs, and 10 training epochs. All runs were done on a CPU without a GPU. The best model was chosen using the macro-averaged F1 score on a separate validation set, and that saved checkpoint was then used to report results on the unseen test data.

## 3. Results

### 3.1 Sentiment Classification Results

For the full model trained on 300,000 graphs, the confusion matrix shows that most sentences are correctly assigned to the three sentiment classes. For negative sentences, 1,661 are predicted as negative, while 436 are softened to neutral and 204 are wrongly marked as positive, so the model is strongest at detecting negatives but sometimes shifts them towards neutral. For neutral sentences, 9,238 are correctly predicted as neutral, with 215 pushed to negative and 987 to positive, which suggests the model usually recognises neutral tone but can be pulled towards neighbouring classes when the wording is a bit emotional. For positive sentences, 15,698 are predicted as positive, 1,382 as neutral, and only 179 as negative, meaning positive feedback is captured very well and is rarely flipped into the opposite class.

When the same GAT model is trained on a smaller set of 50,000 graphs, the confusion matrix shows a similar pattern, just with fewer examples. Among negative sentences, 309 are predicted correctly, with 140 moved to neutral and 70 to positive, again showing that some negative tone is softened to neutral. For neutral sentences, 2,506 are predicted as neutral, 49 as negative, and 322 as positive, while for positive sentences, 5,904 are correctly identified, 619 are shifted to neutral, and only 81 are misclassified as negative. Overall, comparing the two matrices shows that performance improves as the training size increases from 50k to 300k graphs: the number of correct predictions on the main diagonal rises, and the share of serious mistakes, such as confusing positive with negative, stays low and is smallest in the 300k case. Figure 5 shows the confusion matrix for GAT Model.

**Figure 5.**
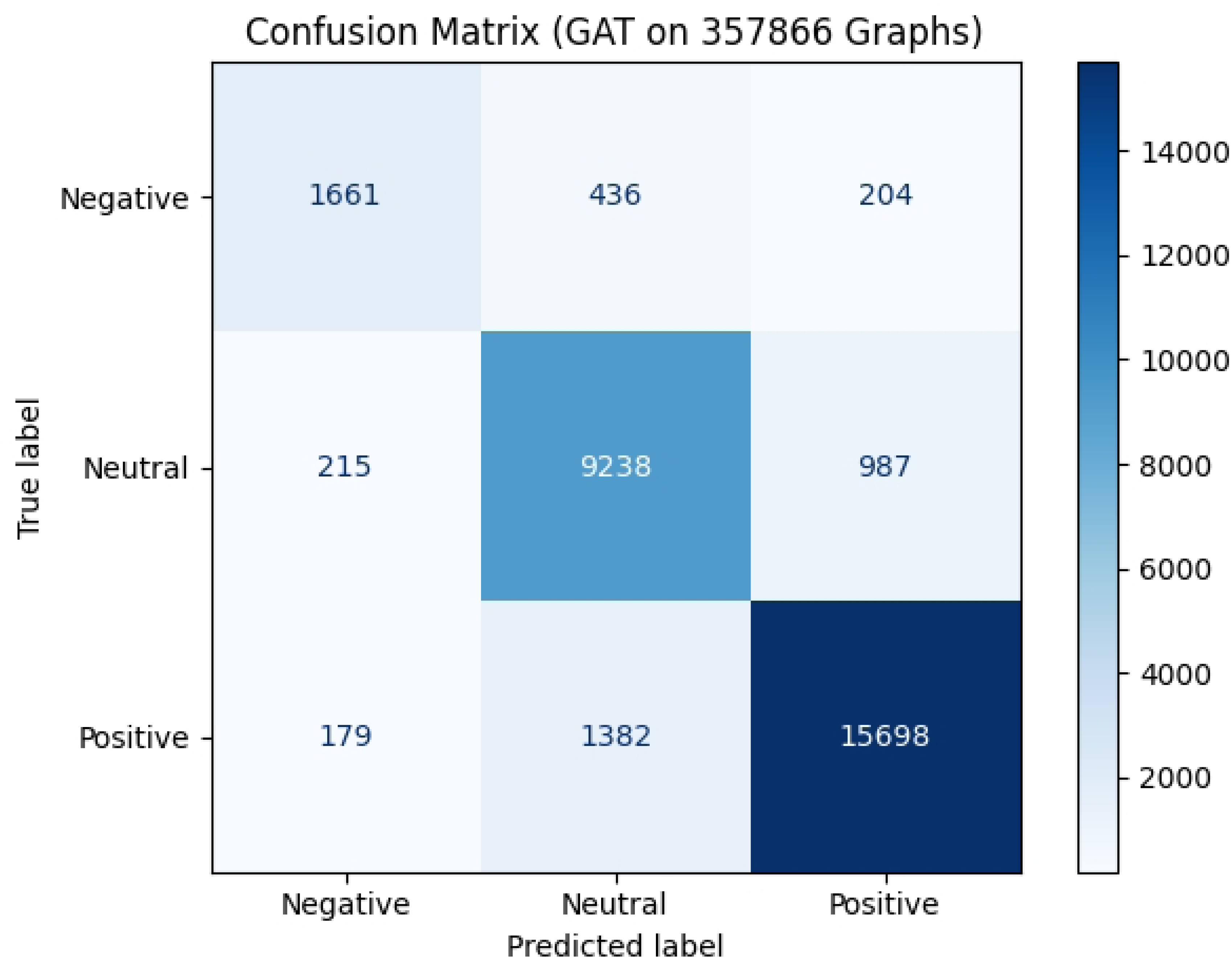
Confusion matrices for sentiment classification on training data. (A) Model trained on 50,000 graphs. (B) Model trained on 300,000 graphs. The matrices show predicted versus actual sentiment labels (negative, neutral, positive) for aspect-level sentiment classification.

#### 3.1.1 Baseline Comparison

Table 3 presents a comparison of the proposed GNN-ABSA framework with commonly used baseline sentiment analysis approaches.

**Table 3:** Comparison of Proposed Model with Baseline Sentiment Analysis Approaches.

| Method | Aspect-Level Modelling | Syntactic Dependency Modelling | Explainability | Typical Limitations | Representative Studies |
| --- | --- | --- | --- | --- | --- |
| Lexicon-based (VADER) | No | No | High | Domain-insensitive, limited handling of context and negation | Hutto and Gilbert [9] |
| Traditional ML (SVM, LR) | Limited | No | Medium | Requires manual feature engineering; weak performance on complex language | Fang *et al.* [5] |
| BERT-based Models | Yes | No | Medium | Sequence-based; limited explicit modeling of word dependencies | Devlin *et al.* [3] |
| Dependency-based ABSA | Yes | Partial | Medium | Relies on predefined dependency rules; scalability issues | Sun *et al.* [19] |
| Proposed GNN-ABSA | Yes | Yes | High | Higher training cost; graph construction overhead | This study; Alhadlaq and Altheneyan [1] |

#### 3.1.2 Use of VADER in Health Application Sentiment Analysis

Lexicon-based sentiment analysis methods, especially the Valence Aware Dictionary and sEntiment Reasoner (VADER), are often used as baseline tools for health-related user reviews because they are simple to apply, easy to interpret, and require little computing power [9]. VADER is often used in work with app store reviews and social media data when researchers need quick sentiment estimates and want to avoid time-consuming model training.

In studies of health and mobile health (mHealth) apps, VADER has been used to analyse user feedback to track overall sentiment patterns and levels of user satisfaction. For instance, Fang et al. applied a lexicon-based sentiment approach to large-scale app review data, showing that it can serve as a lightweight baseline, even though it has clear limits with domain-specific language and context-dependent meaning [5].

Similarly, earlier work on mining mobile app reviews has often used lexicon-based sentiment methods to get an initial view of sentiment distribution before applying more advanced machine learning or deep learning models [12], [16].

However, many studies also note that lexicon-based methods such as VADER have difficulty handling more complex language features, including negation scope, sarcasm, and aspect-level sentiment, especially in noisy, domain-specific text like health app reviews [9], [16].

These limitations have led researchers to turn to more advanced models that use context and sentence structure, such as graph-based neural networks, to carry out more detailed sentiment analysis in health app settings.

#### 3.1.3 Use of VADER in Sentiment Analysis

Table 4 summarizes the use of VADER in health and mobile app sentiment analysis studies.

**Table 4.**
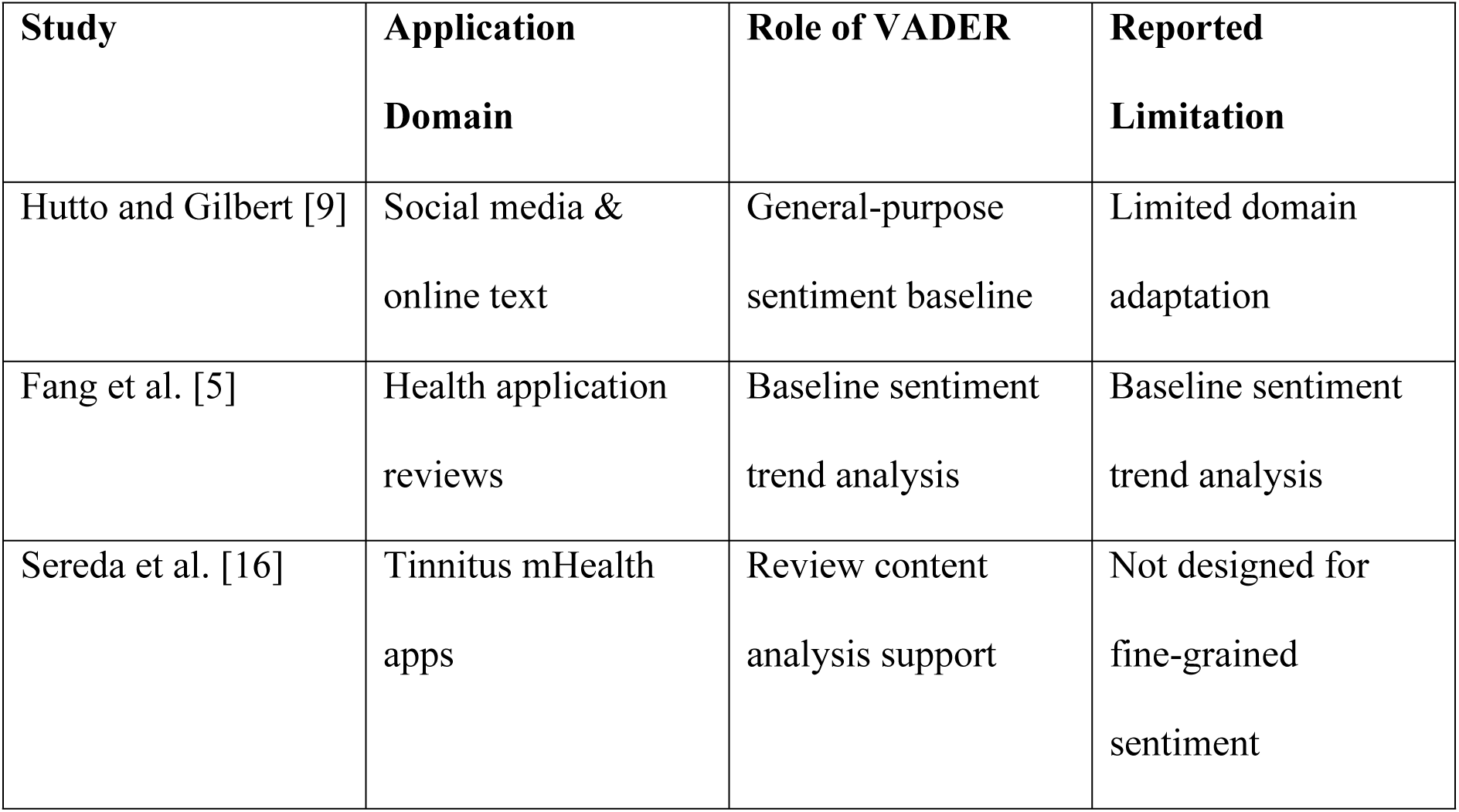
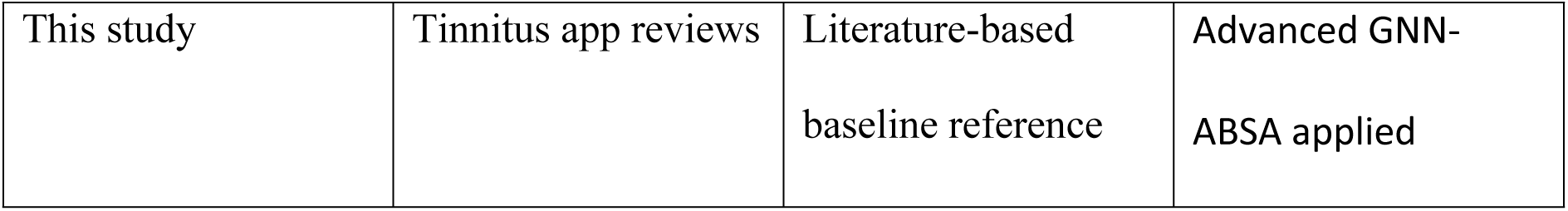
Use of VADER in Health and Mobile App Sentiment Analysis.

### 3.2 Evaluation on Unseen Data

On the unseen test split, the model was evaluated on 57,864 graphs that were not used during training. On this data, it achieved an accuracy of 0.844 and a macro-F1 score of 0.829, which shows that performance stayed strong on new examples.

The confusion matrix for these 57k graphs follows a clear pattern. For negative cases, 9,419 are predicted as negative, 2,562 as neutral, and 553 as positive. For neutral sentences, 27,124 are predicted as neutral, while 1,141 are shifted to negative and 1,112 to positive. For positive sentences, 12,292 are predicted as positive, 2,911 as neutral, and only 750 as negative. Most of the values sit on the main diagonal of the matrix, which means the model keeps a similar structure of predictions to the training phase and generalizes well to unseen graphs. “Figure 6 and Table 5 show the confusion matrix and accuracy results.“

**Figure 6.**
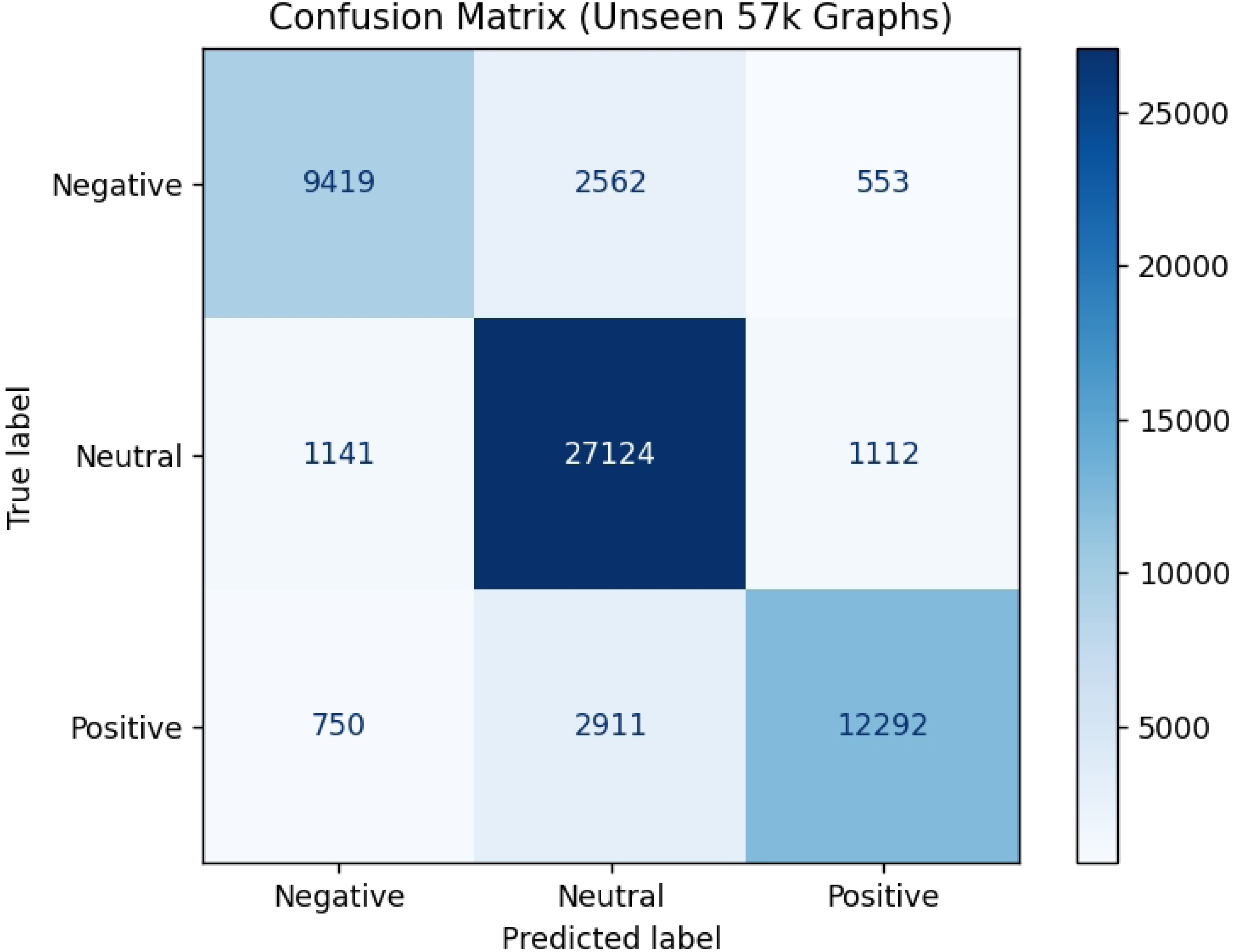
Confusion matrix for sentiment classification on unseen test data (57,864 graphs). The matrix demonstrates model generalization performance on previously unseen aspect-sentiment pairs, with 84.4% overall accuracy.

**Table 5.** Model Performance on Unseen Test Data (Accuracy)

| Unseen Test Results |  |
| --- | --- |
| Accuracy | 0.8440 |
| Macro-F1 | 0.8291 |

The confusion matrix (Figure 6) reveals several important patterns in the model’s classification behaviour. The diagonal values show that the majority of predictions are correct: 76% of negative cases (9,419 out of 12,534), 92% of neutral cases (27,124 out of 29,377), and 77% of positive cases (12,292 out of 15,953) are correctly classified. This indicates the model has learned meaningful patterns from the training data and can generalize to new examples. The most common classification error is the confusion between neutral and positive sentiment, where 1,112 neutral cases are misclassified as positive and 2,911 positive cases are misclassified as neutral. This pattern is understandable given that tinnitus app reviews often express cautious optimism or partial satisfaction, which can blur the boundary between neutral and positive sentiment. For instance, a review stating “it helps a bit but not much” contains both neutral and mildly positive elements. Importantly, the model rarely makes severe errors. Only 553 negative cases are misclassified as positive (4.4%), and only 750 positive cases are misclassified as negative (4.7%). This low rate of sentiment reversal errors indicates the model can reliably distinguish between genuinely positive and genuinely negative user experiences, even if it occasionally struggles with the neutral-positive boundary. The overall accuracy of 84.4% (Figure 7) demonstrates that the GNN-ABSA approach achieves strong performance on aspect-level sentiment classification. This accuracy level is comparable to reported performance in other ABSA studies on product reviews [1] and health app feedback [5], suggesting the graph-based approach generalizes effectively to tinnitus-specific content. The macro-F1 score of 82.9% indicates balanced performance across all three sentiment classes, with no single class dominating the predictions. These results confirm that the model successfully learned to identify aspect-level sentiment patterns from the VADER-labelled training data and can apply these patterns to previously unseen review sentences.

**Figure 7.**
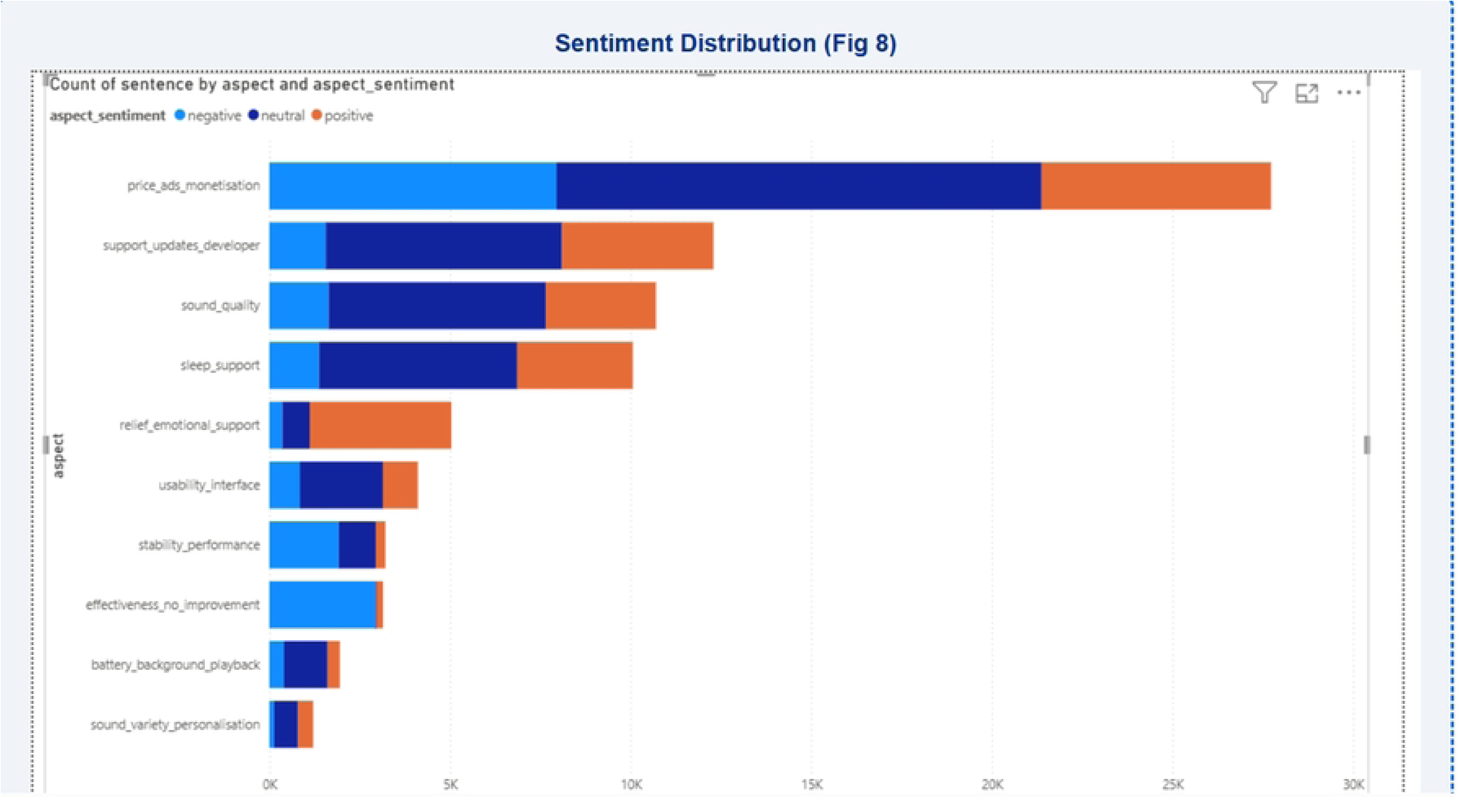
Distribution of sentiment by app feature aspect. The stacked bar chart displays the proportion of positive, neutral, and negative sentiment mentions for each aspect category, revealing feature-specific user satisfaction patterns.

## 4. Discussion

This part of the discussion links the experimental results back to what people with tinnitus actually experience in everyday life. The accuracy numbers matter, but the main value is in what the sentiment patterns say about day-to-day difficulties and ways of coping. By setting these results alongside current evaluation methods, the study can offer more practical and focused suggestions for clinicians and developers who are working on these apps.

### 4.1 Interpretation of Sentiment Patterns

Figure 7 shows that most user comments about tinnitus app features are positive, followed by neutral ones, with negative views appearing less frequently. However, when examined by specific feature, the three sentiment categories reveal different aspects of user experience rather than simply dividing users into satisfied or dissatisfied groups.

Features linked to pricing, adverts, and other monetization choices generate the largest number of sentiment-bearing sentences, and many of these are neutral or negative. This suggests that the way apps charge or show ads is a major source of frustration or uncertainty, even when other parts of the app are viewed more positively. By contrast, therapeutic features such as sound quality, sleep support, and emotional relief more often attract positive comments, which implies that users do value the practical help these features provide, even if the app as a whole is not perfect.

Neutral sentiment shows up a lot across several aspects, including usability, sound, and developer support. This often reflects mixed or conditional feedback, where users find the app helpful in some situations but still point out issues with the interface, reliability, or cost. The overlap between neutral and positive sentiment seen in the confusion matrices is therefore consistent with genuine ambiguity in how people describe their experiences, rather than being only a modelling error.

Negative sentiment, although less common overall, is heavily focused on particular areas such as pricing, stability, and battery drain. These negative reviews tend to be clear and strongly worded, often signalling that people are close to stopping or have already stopped using the app. The strength of this language helps to explain why negative sentiment is picked up with relatively high precision.

Overall, these patterns suggest that sentiment in tinnitus app reviews covers a spectrum that includes partial relief, cautious optimism, trade-offs, and outright frustration. Looking at sentiment at the level of individual features gives a more realistic view of how people actually use and judge these apps in everyday life.

### 4.2 Feature-Level Insights from GNN-ABSA

A main contribution of this study is the use of aspect-based sentiment analysis together with graph neural networks to capture feature-level user experience at scale. Instead of working at the whole-review level, this approach links sentiment directly to the specific app features mentioned in each sentence, which matters in tinnitus reviews where people often talk about several features with different opinions in a single comment.

By turning each sentence into a separate graph, the model can focus on local context without mixing up unrelated parts of the review. Aspect masking then narrows this down further by making sure the sentiment prediction is driven only by the nodes that match the detected aspect terms. This helps cut out noise from words that are not relevant to that feature and makes the feature-level sentiment more precise.

The analysis shows that user satisfaction is not spread evenly across features. Therapeutic elements such as sound masking and sleep support are more likely to attract positive comments, while design and delivery issues like pricing, adverts, and technical stability are more often linked to negative sentiment. This illustrates how graph-based aspect-level sentiment analysis can reveal practical, feature-specific insights that do not appear in overall star ratings or simple document-level sentiment scores.

### 4.3 Comparison with Prior Evaluation Approaches

Unlike earlier studies that used expert ratings, surveys, or clinical tests, this work takes a user-focused approach based on real-world feedback from over 342,000 reviews. Tools like the Mobile App Rating Scale (MARS) give structured quality scores, but someone has to rate them manually and they miss how people actually feel about specific features day-to-day.

Surveys and focus groups offer useful insights but can’t scale up or capture much diversity. Clinical trials with measures like the Tinnitus Handicap Inventory (THI) or Tinnitus Functional Index (TFI) show therapeutic effects in controlled settings but skip usability, engagement, and whether people keep using apps long-term.

By digging into 342,520 reviews across ten years, this study fills those gaps. Feature-level sentiment analysis uncovers patterns of what users like and dislike that go beyond clinical results or expert views, giving a fuller picture of how tinnitus apps work in everyday life.

### 4.4 Usability of the Research

The main strength of this research is that it can be used in practice. Instead of looking only at overall ratings or general sentiment, it shows how users feel about individual features inside tinnitus apps. This makes the results useful for different groups who work with these tools.

For app developers, the findings point to specific features that need attention, such as background playback, adverts, subscription handling, and technical stability. Focusing on these areas can help them priorities changes that are most likely to improve user satisfaction and keep people using the app over time.

For clinicians, the results give a clearer picture of how patients experience tinnitus apps in everyday life, outside formal trials. Clinical studies may show that an app can help, but this work helps explain why people sometimes stop using it. That can support more realistic recommendations and better management of patient expectations.

For people with tinnitus, the study suggests that it is better to choose apps based on specific needs—such as sleep support, stress relief, or hearing-aid compatibility—rather than relying only on overall star ratings.

### 4.5 Connection to Explainable Artificial Intelligence

This study relates closely to the principles of Explainable Artificial Intelligence (XAI). The GNN-based model represents each review as a graph that shows how words in a sentence are connected, allowing emotional expressions to link directly to the features they describe.

Unlike black-box sentiment classifiers, the proposed approach supports transparency by linking sentiment predictions to specific app features, aligning with established principles of explainable artificial intelligence [29], [30]. For instance, negative feedback about background playback often includes words like “stops,” “crashes,” or “screen locks.” Transparency is especially valuable in healthcare applications, where users and professionals need to understand how a system reaches its conclusions. By explaining results at the feature level, the model supports both technical insight and practical improvement of tinnitus apps.

### 4.6 Feature-Level Insights Beyond Positive, Negative, and Neutral

A key contribution of this research is moving beyond simple positive, negative, or neutral sentiment labels. Many reviews express mixed or conditional experiences that cannot be captured by a single overall sentiment score. Beyond identifying which features attract positive or negative sentiment, the feature-level analysis also reveals how users describe their experiences in ways that do not fit neatly into a single sentiment category. Many reviews contain language that reflects trade-offs, uncertainty, or conditional satisfaction. For example, users often acknowledge that sound therapy helps them relax or sleep, while simultaneously reporting frustration when playback stops, ads interrupt sessions, or the app crashes during use. In such cases, a simple positive or negative label would hide the underlying reason for dissatisfaction. Neutral sentiment, in particular, plays an important role in understanding user experience. Rather than indicating indifference, neutral reviews often represent mixed feelings or partial benefit. Users may report that an app “helps a little,” “works sometimes,” or “is good in theory but unreliable in practice.” These comments suggest that users are not rejecting the therapeutic idea itself but are struggling with inconsistent performance or usability barriers. This helps explain why neutral sentiment appears frequently across several features, especially usability, stability, and sleep-related aspects. Negative sentiment tends to be more focused and direct. Strongly negative reviews often mention repeated errors, subscription problems, background playback failures, or compatibility issues with hearing aids and newer operating systems. These comments frequently include expressions of abandonment, such as uninstalling the app shortly after installation. From a practical perspective, this indicates that negative sentiment is closely tied to points where users disengage entirely, rather than to dissatisfaction with the therapeutic concept.

Using feature-level GNN analysis, clear and consistent patterns emerge:

- **Sound therapy → Positive** Users often report that sound masking and noise options help with relaxation and sleep.
- **Sleep support → Mixed** Benefits are frequently reported but depend on uninterrupted playback.
- **Advertisements → Negative** Ads are widely described as disruptive, especially during sleep.
- **Background playback → Negative** Many users report that audio stops when the screen is locked, which directly undermines the app’s purpose.
- **App stability → Negative** Crashes and repeated error messages lead to frustration and quick uninstallation.
- **Pricing and subscriptions → Negative** Transaction failures and paywalls discourage continued use.

Taken together, these patterns show that sentiment in tinnitus app reviews reflects a spectrum of lived experience rather than a simple like–dislike response. Feature-level GNN analysis makes it possible to separate therapeutic value from delivery problems, revealing that many users are willing to engage with sound therapy but are discouraged by avoidable design and technical issues. This deeper interpretation helps explain why some apps receive reasonable overall ratings yet still suffer from poor long-term retention. The findings suggest that addressing implementation failures—such as improving background playback reliability, reducing intrusive advertising, and enhancing app stability—could substantially improve user satisfaction without requiring changes to the underlying therapeutic approach.

### 4.7 Helping Users Find the Right App and Market Implications

This research can also help people choose tinnitus management apps that fit their own needs. Rather than looking only at overall star ratings, users can pay attention to feedback on specific features, such as how well an app supports background playback or how reliably it works with hearing aids.

From a market point of view, the results help explain why many apps fail to keep users over time. A lot of the negative feedback is caused by design and delivery problems that could be avoided, rather than by the basic therapeutic idea. Fixing issues like pricing, adverts, or stability can improve user retention, app ratings, and trust in digital health tools.

This way of looking at the data is useful for developers and companies who want to increase both user satisfaction and commercial success.

### 4.8 Readability, Relevance, and Broader Appeal

This paper is written so it can be read by a broad audience, including computer science researchers, digital health professionals, clinicians, and app developers. Using real-world data from more than 300,000 reviews makes the work relevant outside tightly controlled lab or trial settings.

By bringing together graph-based machine learning and practical user feedback, the study links technical methods with real-life impact. The use of clear examples and concrete implications is intended to make the results easy to follow and useful for readers from different backgrounds.

### 4.9 Limitations

While this study demonstrates that large-scale, graph-based aspect-level sentiment analysis can support the evaluation of tinnitus management apps, it also has some limitations that should be acknowledged.

First, the analysis uses only reviews from public app stores. These comments reflect the experiences of many users but don’t represent everyone who uses the apps. People who post reviews usually have very strong opinions—either positive or negative—so those with moderate or brief experiences are likely underrepresented.

App store reviews also lack personal details such as age, tinnitus severity, symptom duration, or other health conditions, all of which may influence user experiences and expectations. Because of this, the results reflect users’ perceptions of the apps rather than their measured clinical outcomes.

Second, the sentiment labels were generated automatically rather than assigned by human reviewers. Although the graph neural network performed consistently on new data, automated methods can still produce small errors. This matters in tinnitus-related discussions, where people often share mixed or uncertain feelings instead of clearly positive or negative ones. Previous research notes that analysing short, informal comments can make it difficult to capture this subtlety, especially in health contexts [26]. As a result, some emotional nuances may not be fully reflected within a simple three-class sentiment system.

Third, the aspect extraction used a fixed list of features specific to the tinnitus domain. These aspects were chosen to reflect key parts of the apps, but they don’t cover every possible element of user experience. Some reviewers mention personal coping methods or how the app connects with other devices, which fall outside the set categories. Recent studies on mobile health assessment suggest that more adaptive, data-driven approaches could help identify new features as apps continue to evolve [14].

Fourth, the graph design used sentence-level representations to keep enough context while keeping the computation manageable. This approach made it possible to analyse hundreds of thousands of reviews, but some links between sentences in longer comments may have been lost. Many users describe their experiences across several sentences, building context gradually. Using document-level or hierarchical graph models might reveal more of these patterns, though they would need far greater computing resources. This highlights an ongoing issue in using graph neural networks with healthcare data—finding a workable balance between scale and interpretability [13],[2].

Fifth, all experiments were carried out on a local computer using CPU-based training. This setup made the work more practical and easier to run, but it also meant that more complex models, such as deeper graph networks or extensive hyperparameter tuning, couldn’t be tested. Previous studies on graph neural networks in healthcare point out that hardware limits and time constraints often affect which models can be used and may restrict how much performance can be improved [13].

Finally, this study considered only English-language reviews from major app stores. Tinnitus affects people in many countries, so experiences shared in other languages or cultural contexts were not included. Factors such as healthcare access, availability of apps, and local attitudes toward digital health may shape how people rate or use tinnitus apps. Expanding the analysis to multilingual data could make the results more representative and support broader international comparisons of tinnitus-related mobile health tools [24].

Even with these limitations, the study offers a large-scale, user-focused view that complements clinical and expert assessments. The findings also highlight areas for further research, such as improving the interpretability of graph-based models, using more flexible methods for aspect detection, expanding sentiment analysis to multiple languages, and exploring how user feedback relates to clinical outcomes.

## 5. Conclusion

This study examines user opinions on tinnitus management apps by analysing over 342,520 app store reviews with a graph-based, feature-level sentiment method. Combining graph neural networks with aspect-based sentiment analysis provides a deeper view of how users respond to individual app features rather than relying only on overall star ratings.

Working with reviews from 84 tinnitus apps collected over ten years, the analysis ran smoothly on standard hardware. The results show sentiment varies widely across features. Therapeutic tools like sound masking and sleep support mostly get positive comments, though these tend to reflect partial relief rather than full symptom relief. By contrast, pricing, adverts, and usability problems consistently draw neutral or negative feedback. This suggests user frustration often comes from fixable design decisions rather than the core therapeutic idea.

What stands out is how sentiment in these reviews captures broader patterns of living with tinnitus, not just simple like/dislike reactions. Positive comments often mix cautious hope with practical coping, neutral ones show trade-offs and mixed feelings, and negative ones point to clear frustration or reasons for quitting the app. This explains patterns in the classification results, like neutral bleeding into positive, and shows why feature-level analysis matters for chronic, personal conditions.

Methodologically, turning sentences into graphs with aspect masking made it possible to link sentiment precisely to individual features. Unlike expert ratings, surveys, or clinical measures, this approach uses large-scale real-world feedback to capture what users actually say about specific app elements—insights that controlled studies and expert reviews often miss.

The work also points to clear next steps. More flexible aspect discovery could adapt to new app features over time. Multilingual analysis would catch cultural differences in app use. Explainable GNN techniques would build trust in healthcare settings. And combining sentiment patterns with clinical data like tinnitus severity scores would show how user experience ties to actual treatment outcomes.

In summary, large-scale graph-based feature sentiment analysis offers a practical way to understand tinnitus apps. It captures real user experience both broadly and at feature level, complementing clinical trials and expert evaluations while giving developers, clinicians, and researchers concrete insights to work with. As digital health tools grow, this kind of user-focused computational analysis will be key to creating apps that deliver in practice, not just look promising on paper.

## Data Availability

The datasets analysed during the current study are publicly available in the Zenodo repository at https://doi.org/10.5281/zenodo.18419488. The raw review data were collected from publicly available Apple App Store and Google Play Store platforms. The dataset includes 342,520 cleaned reviews with associated metadata, aspect detection keywords, and comprehensive documentation.

## Abbreviations

ABSA: Aspect-Based Sentiment Analysis
API: Application Programming Interface
BERT: Bidirectional Encoder Representations from Transformers
CBT: Cognitive Behavioural Therapy
CSV: Comma-Separated Values
EDA: Exploratory Data Analysis
FTRS: Fudan Tinnitus Relieving System
GAT: Graph Attention Network
GNN: Graph Neural Network
MARS: Mobile App Rating Scale
mHealth: Mobile Health
NHS: National Health Service
RCT: Randomized Controlled Trial
TFI: Tinnitus Functional Index
THI: Tinnitus Handicap Inventory
TRT: Tinnitus Retraining Therapy
VADER: Valence Aware Dictionary and sEntiment Reasoner
XAI: Explainable Artificial Intelligence

## Declarations

## Ethics approval and consent to participate

Not applicable. This study analysed publicly available app store reviews and did not involve human participants, human data, or human tissue.

## Consent for publication

Not applicable.

## Authors’ contributions

MNY led the conceptualization and design of the study, performed data collection and preprocessing, implemented the GNN-ABSA model, conducted the statistical analysis, and drafted the manuscript. MNA contributed to the methodological framework, model validation, and critical revision of the manuscript for intellectual content. NN contributed to the sentiment analysis methodology design and interpretation of results. UH contributed to the research design, data analysis framework, and interpretation of findings. All authors participated in refining the research approach, critically reviewed successive drafts, contributed substantive revisions, and approved the final manuscript for submission.

## Acknowledgements

The authors acknowledge Northumbria University for providing open access funding through its institutional agreement with Springer Nature.

